# Soluble IL-2R contributes to impaired muscle cell mitochondrial respiration in fatigued individuals with post-acute sequelae of COVID

**DOI:** 10.1101/2024.08.14.24311980

**Authors:** Laura P. Brown, Jai Joshi, Kate Kosmac, Douglas E. Long, Ashley A. Montgomery-Yates, Anna G. Kalema, Jamie L. Sturgill, Hemendra Vekaria, Patrick Sullivan, Dylan Wilburn, Panagiotis Koutakis, Christine M. Latham, Christopher S. Fry, Philip A. Kern, Benjamin Miller, Esther E. Dupont-Versteegden, Ahmed Ismaeel, Kirby P. Mayer, Yuan Wen

## Abstract

Post-acute sequelae of COVID (PASC) persist in many patients for weeks and months after recovery from initial SARS-CoV-2 infection. Recent evidence suggests that pathological changes in skeletal muscle may contribute significantly to ongoing pain and fatigue, particularly post-exertional malaise. This study aimed to investigate the underlying mechanisms of PASC-related fatigue by examining skeletal muscle function and circulating factors in affected individuals.

We conducted a cross-sectional case-control study of patients with fatigue-associated PASC who had experienced mild to moderate COVID-19 without hospitalization. Skeletal muscle biopsies revealed reduced mitochondrial respiration and content in PASC participants compared to healthy controls. This lower respiratory capacity was accompanied by markedly elevated circulating levels of soluble IL-2 receptor alpha subunit (sIL2R), a T cell-specific receptor.

*In vitro* experiments demonstrated that sIL2R directly impairs mitochondrial oxygen consumption and reduces mitochondrial complex III subunit protein levels in cultured muscle cells. These findings suggest a mechanism linking systemic immune dysregulation to muscle-specific mitochondrial dysfunction in PASC.

This work provides new insights into the pathophysiology of PASC identifying sIL2R as a promising therapeutic target for addressing mitochondrial deficits in PASC-related fatigue and opening avenues for developing targeted interventions.

## BACKGROUND

Coronavirus disease 2019 (COVID-19), caused by the respiratory virus SARS-CoV-2, presents with a wide spectrum of symptoms ranging from mild flu-like illness to severe, life-threatening conditions [1–3]. While many individuals recover fully from acute infection, a significant proportion of patients experience persistent or newly emerging symptoms lasting months after initial recovery [4–8]. This condition is defined as persistent and/or delayed complications or long-term complications of SARS-CoV-2 infection beyond four weeks from the onset of symptoms [4, 9]. Common symptoms include dyspnea, joint pain, headaches, brain-fog, muscle weakness and fatigue [4–7, 10].

Prolonged fatigue stands out as one of the most prevalent and debilitating complaints in patients with PASC. This fatigue is often accompanied by unexplained muscle and joint pain, suggesting a potential link to underlying musculoskeletal pathology [8, 11]. Recent studies have associated PASC fatigue with structural and functional mitochondrial abnormalities in skeletal muscle [12, 13] , pointing towards a possible mechanistic explanation for the persistent symptoms. Notably, PASC patients exhibit elevated T cell levels and dysregulation of both humoral and cellular immune responses [14]. This immune dysregulation may play a crucial role in the development and persistence of PASC symptoms, particularly in skeletal muscle tissue as a key site of pathology in post-COVID fatigue. Three recent studies point to an elevated inflammatory response, mediated by T lymphocytes both locally within muscle tissue and systemically, and may contribute to impaired muscle function and reduced mitochondrial respiration [12, 13, 15].

The observed deficits in mitochondrial respiration and muscle performance in PASC patients, coupled with prolonged immunological changes, led us to hypothesize that a humoral response to SARS-CoV-2 is the primary cause of skeletal muscle pathology. Specifically, we suggest that certain circulating factors, most likely T-cell derived, are elevated in PASC and induce mitochondrial abnormalities [16–22]. To test this hypothesis, we conducted an analysis of plasma immune markers of PASC participants to identify candidates that may directly or indirectly impair muscle cell mitochondrial respiratory capacity. Our findings revealed significant reductions in measures of mitochondrial content and impaired muscle energetics in individuals suffering from fatigue specific PASC. We identified the T cell-derived soluble IL-2 receptor alpha subunit (sIL2R) as the most significantly elevated systemic factor following recovery from SARS-CoV-2 infection.

## METHODS

### Study Design and Participants

This prospective, cross-sectional study was approved by the University of Kentucky’s institutional review board (MEDFL 42971,43046, 43499, 46072). Participants must be 18 years or older and provided informed consent, and the study adhered to the Helsinki Declaration. Inclusion criteria for PASC participants were defined as follows: individuals who reported a positive RT-PCR test or at home antigen test for SARS-COV-2, reported “mild to moderate illness” and experienced post-COVID fatigue at least 4 weeks after positive COVID test were eligible. Mild to moderate illness was defined as having at least one sign or symptom of COVID but not requiring supplemental oxygen or hospitalization [1]. Participants reporting hospitalization in the previous 12 months or any time during the SARS-COV-2 positive period until the date of research testing were excluded. Subjects were recruited to participate in a comprehensive assessment of muscle and physical function, and muscle biopsy and blood collection at least 4 weeks after infection. Due to the nature of SARS-COV-2 and rapid transmission, to guarantee COVID-negativity, we used historic convenience biopsy samples collected pre-pandemic and stored in the University of Kentucky Center for Muscle Biology Skeletal Muscle Biobank, as healthy controls when possible. For healthy control HRR experiments and immune markers, historic convenience samples from pre-pandemic as well as COVID negative pandemic samples were chosen with the same inclusion criteria [23, 24]. Age for healthy participants (n=25, 40% female) was 28.0±14.0 years and for PASC subjects (n=11, 45.5% female) was 32±21 years. BMI for healthy participants was 26.3±6.2 kg/m^2^ and for PASC subjects was 27.13±1.7 kg/m^2^. Age and BMI are reported as median and interquartile range.

### Blood collection and muscle biopsy

Vastus lateralis muscle biopsies were obtained using a 5 mm Bergstrom needle as previously described [24]. Blood samples were collected for serum and plasma analysis. A subset of participants (n=7 PASC, n=9 control) consented to consume deuterium oxide for 7 days prior to biopsy to assess protein synthesis.

### Histology and Microscopy

Muscle biopsies were sectioned at –22 to –25°C and stained for succinate dehydrogenase (SDH) prior to immunohistochemistry. Incubation with primary antibody cocktail was performed overnight at room temperature: Mouse IgG2b BA.D5 concentrate (1:100, Type 1 MyHC), Mouse IgG1 SC.71 supernatant (1:50, Type 2a MyHC), Mouse IgM 6H.1 supernatant (as diluent, Type 2x MyHC) and rabbit anti-laminin to stain fiber borders. Secondary antibodies were applied for 1 hour at room temperature: anti-mouse IgG2b AlexaFluor 647, IgG1 AlexaFluor 488 and IgM AlexaFluor 555 (Thermo Fisher) and anti-rabbit biotin (Jackson ImmunoResearch). Finally, sections were incubated with streptavidin-AMCA (Vector Laboratories), imaged with a 20x objective using an Olympus BX61 VS microscope, and analyzed with MyoVision analytical software to determine fiber cross-sectional area (CSA) and fiber type frequency [25]. All type 2a and 2x fibers were combined for analysis. SDH total fiber type specific categorization of light, intermediate, and dark fibers was carried out by two blinded researchers using threshold intensity in image J.

### Muscle and Physical Function Assessment

Participants underwent: 1) Timed-up and Go (TUG) test to assess functional mobility and TUG-cognitive test to assess dual-task performance during a mobility challenge; slower times indicating worse performance [26]; 2) Six-minute walk test (6-MWT) was performed with the ATS guidelines to examine functional exercise capacity, and the percent achieved of the expected distance was calculated [27]; 3) Knee extensor strength assessment using hand-held dynamometry (Lafayette Manual Muscle Test System Model-01165, Lafayette Company, Lafayette, IN) in supine position with knee on a bolster in 20-30⁰ of flexion.

### Physical Activity Survey

Participants were asked three questions: the level of their physical activity prior to SARS-CoV-2 infection, whether their physical activities had reduced, and if they have suffered from myalgias since recovery from acute symptoms.

### Ultrasonography

Ultrasound imaging of quadriceps musculature was performed to assess muscle thickness. Ultrasound B-mode images of quadriceps musculature (rectus femoris, vastus intermedius) were acquired using Sonosite IViz ultrasound system as previously described [28–30].

Quadriceps complex muscle thickness is measured between the uppermost part of the bone echo of the femur and the superficial fascia of the rectus includes rectus femoris and vastus intermedius muscles. Rectus femoris thickness is measured between 2 transverse fascial planes of the belly of the rectus femoris muscle utilizing the center of the femur as a marker to maintain orientation.

### Mitochondrial Analyses

#### High-resolution respirometry (HRR)

Permeabilized muscle fiber bundles were prepared from freshly isolated biopsies of the vastus lateralis muscle in both PASC and healthy participants (n = 10 healthy, 7 PASC). To assess mitochondrial respiration, 15 to 20 milligrams of fresh tissue were placed into BIOPS solution for HRR using an O2k fluorespirometer (Oroboros Instruments, Innsbruck, Austria). Connective tissue was removed, and samples were gently tweezed apart to separate fiber bundles. Fiber bundles were then permeabilized in BIOPS solution with 50 μg/ml saponin for 30 min at 4°C. Following permeabilization, samples were washed in MiR05 mitochondrial respiration medium (0.5 mM EGTA, 3 mM MgCl2, 60 mM lactobionic acid, 20 mM taurine, 10 mM KH2PO4, 20 mM HEPES, 110 mM D-sucrose, 1 g/L fatty acid free Bovine Serum Albumin) for 10 min at 4°C, and then roughly 2 mg of permeabilized fibers were added to the O2k chamber in a total volume of 2 ml of MiR05, supplemented with 20 mM creatine. HRR was determined at 37°C in hyperoxic conditions (220-450 μM O_2_) using a substrate-uncoupler-inhibitor titration (SUIT) protocol using 5 mM pyruvate + 2 mM malate (to support electron flow through complex I), 2.5 mM ADP (to stimulate oxidative phosphorylation (OXPHOS)), 10 μM cytochrome c (to assess outer mitochondrial membrane integrity, defined as <15% increase), 10 mM glutamate (additional complex I substrate), 10 mM succinate (to support electron flow through complex II), 0.5 μM titrations of carbonyl cyanide 3-chlorophenylhydrazone (CCCP) (to assess maximum electron transfer capacity), and finally, 0.5 μM rotenone to inhibit complex I, and 2.5 μM antimycin A to inhibit complex III to measure maximal electron transfer capacity of complex II and residual oxygen flux, respectively. Respiratory fluxes were normalized to tissue wet weight added to the O2k chamber.

#### Citrate synthase activity

Citrate synthase (CS) activity was used to estimate mitochondrial content, as previously described [31]. Briefly, skeletal muscle samples were homogenized by glass pestle in RIPA lysis buffer (50 mM Tris-HCl, 150 mM NaCl, 1.0% NP-40, 0.5% sodium deoxycholate, 1.0 mM EDTA, 0.1% SDS, 0.01% sodium azide, pH 7.4). Assays were performed in buffer consisting of sample protein lysate, 100 mM Tris (pH 8.0), 10 mM 5,5′-Dithiobis-(2-nitrobenzoic acid) (DTNB), 30 mM acetyl CoA, and 10 mM oxaloacetic acid (OAA). CS activity was determined by monitoring the change in absorbance at 412 nm using a Cytation 5 Multimode reader (Biotek). All samples were assayed in duplicate in 96 well plates. Activity was normalized to sample protein concentrations, determined using a Pierce BCA Protein Assay (Thermo Fisher Scientific, #23225).

#### Cytochrome c oxidase activity

For measurement of Cytochrome C oxidase (CCO) enzymatic activity (Complex IV of the electron transport chain), the rate of disappearance (oxidation) of reduced cytochrome c was monitored using a CCO assay kit, according to manufacturer’s instructions (Sigma-Aldrich, CYTOCOX1). Skeletal muscle or cell culture lysates homogenized in RIPA buffer were diluted in enzyme dilution buffer. Assay reaction buffer contained sample protein lysate and reduced (ferro)cytochrome c (2 mM, prepared using 0.1 M Dithiothreitol (DTT), for a final concentration of 0.5 mM DTT). CCO activity was determined by monitoring the absorbance at 550 nm for 10 min using a Cytation 5 Multimode reader. All samples were assayed in duplicate, and activity was normalized to sample protein concentrations.

#### Western analysis

Samples lysed in RIPA buffer, as described above, were prepared in Laemmli sample buffer (Bio-Rad Laboratories, Hercules, CA, # 1610737), and twenty micrograms of whole cell homogenate was separated by SDS-PAGE using 4-20% Criterion TGX Stain-Free Protein Gels (Bio-Rad #5678094) at 200V for 1 hour. Total protein stain was activated by brief ultraviolet (UV) excitation using a ChemiDoc MP System (Bio-Rad). The protein was then transferred to a PVDF membrane for 1 hour 30 minutes at 200 mA at 4°C, blocked with 5% milk in TBS-T for 1 hour, and blotted at a 1:200 dilution with anti-OxPhos Human Antibody cocktail (Thermo Fisher #45–8199) for human muscle lysates or anti-OxPhos Rodent Antibody cocktail (Thermo Fisher # 45–8099) for C2C12 lysates in 5% milk in TBS-T overnight at 4°C. The membrane was washed three times with TBS-T before being incubated for 1 hour with goat anti-mouse IgG, HRP-conjugated secondary antibody (1:10,000) (Thermo Fisher # 31430). Blots were developed with Radiance ECL substrates (Azure Biosystems # AC2204). After detection of OXPHOS proteins, the membrane that includes tubulin was blotted with 1:2,000 rat anti-mouse α-tubulin IgG antibody (Thermo Fisher #MA1-80017) for 1 hour at room temperature, washed as described above, and then blotted with goat anti-rat 680nm Alexa Fluor IgG (Thermo Fisher #A-21096) at a dilution of 1:20,000 for 1 hour. Images were acquired using a ChemiDoc MP system (Bio-Rad), and quantified using either Image Lab Software (Bio-Rad) or ImageJ [32].

#### Mitochondrial protein synthesis

Subjects consumed deuterium oxide (D_2_O, 70%, Sigma-Aldrich) starting with a bolus dose (50 ml, 3x/day) followed by 50 ml 2x/day for the 7 days leading into the muscle biopsy. This strategy achieved an isotopic steady state of 1%–2%, as confirmed by a blood draw on the biopsy date, to label newly synthesized skeletal muscle protein [24, 33–35] On the biopsy date 50 mg of tissue was snap frozen for subsequent analysis. For analysis of muscle mitochondria protein synthesis, tissues were fractionated to isolate mitochondria and water enrichment was assessed as previously described [36]. The fraction of proteins that was newly synthesized was calculated from the enrichment of alanine bound in mitochondrial proteins over the entire labelling period, and then divided by the precursor enrichment, using plasma D2O enrichment with Mass Isotopomer Distribution Analysis adjustment.

#### Transmission Electron Microscopy (TEM)

Muscle samples (3-5 mg) from the vastus lateralis were fixed in 2.5% glutaraldehyde 0.1 M sodium phosphate buffer (pH 7.2) solution at 4°C for 48 hrs. The samples were then washed 3 times for 10 minutes in phosphate buffer solution before secondary fixation in 1% osmium tetroxide (OsO4) for 2 hrs. Samples were rinsed three times, for 10 minutes in phosphate buffer solution. Samples were then dehydrated using sequentially increasing concentrations of acetone (50%, 70%, 90%, and 100%) 2 times 10 minutes at each concentration. Samples were embedded with increasing gradients of Embed812:Acetone (1:2, 1:1, 2:1) with a final immersion in 100% Embed 812 (Electron Microscopy Sciences Catalog #70,020, Hatfield, PA, USA) before being polymerized for 48hrs at 60°C. Ultrathin sections were cut at 70nm thickness with a Leica Ultracut EM UC7 ultramicrotome (Leica Microsystems, Wetzlar, Germany), and post-section staining was completed by incubating grids with 2% uranyl acetate for 15 min and 1% lead citrate for 5 min. Prepared samples were analyzed using a JEM1010 transmission electron microscope (JEOL, Tokyo, Japan).

### Analysis of Plasma Immune Markers

The ProcartaPlex Human Immune Monitoring Panel, 65plex (EPX650-10065-901, Thermo Fisher Scientific, Waltham, MA, USA) was used to assess 65 cytokine, chemokine, growth factor, and soluble receptor targets simultaneously using Luminex xMAP technology. Twenty-five microliters of serum or plasma from each sample were processed in duplicate according to manufacturer’s instructions.

#### In Vitro studies

C_2_C_12_ Cells were cultured in growth media (DMEM supplemented with 10% fetal bovine serum, CellPro™, Alkali Scientific, #DMEM15) to confluence and then differentiated (DMEM supplemented with 2% heat inactivated horse serum for 48 hours in differentiation media before the addition of sIL2R (2 ng/ml) or IL-2 (35 ng/ml) in differentiation media. Concentrations of IL-2 or sIL2R were chosen to reflect the average plasma concentration from the PASC cohort.

Twenty-four hours after treatment, oxygen consumption rate (OCR) was measured using the Seahorse XF Cell Mito Stress Test Kit (Agilent #103015-100) and Seahorse XF24 Analyzer (Agilent). Prior to OCR analysis, cell culture media was replaced with Seahorse XF DMEM media (Agilent #103680-100) supplemented with 10 mM glucose, 1 mM pyruvate, and 2 mM glutamine. Compounds injected into the wells and their respective concentrations were as follows: 1.5 μM oligomycin (for inhibition of ATP synthase and measurement of ATP-linked OCR), 1 μM carbonyl cyanide-p-trifluoromethoxyphenylhydrazone (FCCP, for uncoupling of oxidative phosphorylation and measurement of maximal OCR), and 0.5 μM antimycin A/rotenone (for inhibition of Complexes I and III and measurement of nonmitochondrial OCR).

Immediately after OCR measurement, XF DMEM media was removed, and RIPA buffer was added to lyse the cells and determine protein concentrations using a Pierce BCA Protein Assay (Thermo Fisher Scientific #23252). Western blot analysis was performed as described above.

#### Statistical Analysis

Values are presented as means ± standard deviation (SD). In human data, all individual points represent a participant; for *in vitro* data, all individual points represent a replicate. Continuous parametric data was analyzed using Student T-Tests (and a Welch correction when appropriate) or Two-way ANOVA with Tukey post-hoc testing. Continuous non-parametric data were analyzed using a Mann-Whitney *U* tests, or Multiple Mann-Whitney *U* tests. Distributions were assessed with Sharpiro-Wilk tests and variance was assessed using F-tests. Significance was defined as *p* < 0.05 and trending was defined as *p* > 0.05 ≤ 0.1.

## RESULTS

### PASC participants report reduced physical activity, but fatigue is not correlated with compromised muscle mass and contractility

Answers to physical activity prior to CoV-2 infection and current symptom questions revealed that 18% reported high activity levels (4 to 6 days of intense exercise), 36% reported intermediate activity levels (3 to 5 days of exercise) and 45% reported low activity levels (occasional walks, yoga, or activities of daily living, etc.) prior to infection (**Table 1**). Seventy-three percent of participants (n = 8) reported reduced activity since initial infection; the average time since positive test was 59±24 days (average ± standard deviation, **Table 1**). Three participants (27%) reported no change in activity after infection, of which two had been highly active, and one was intermediately active prior to infection.

**Table 1.** PASC participant self reported characteristics. Days since positive test is reported as average days and standard deviation. Activity level prior to infection, change in activity, and chronic myalgia since infection is reported as quantity reporting and percentage of all PASC participants who answered (n=11).

| COVID participant characteristics | Results |  |
| --- | --- | --- |
| Days since positive SARS-CoV-2 test | 59.36 ±24.44 days |  |
| Activity level prior to infection | High | 2(18.18%) |
|  | Intermediate | 4(36.36%) |
|  | Low | 5(45.45%) |
| Change in activity | Reduced | 8 (72.73%) |
|  | No change | 3 (27.27%) |
| Chronic myalgia since infection | Yes | 3 (27.27%) |
| *Inclusion criteria for PASC included subjective report of fatigue at time of enrollment and biopsy testing.* |  |  |

We assessed healthy and PASC participants for their ability to perform timed-up and go (TUG) tests both with (**Fig 1A**) and without (**Fig 1B**) simultaneous cognitive testing and found no significant differences between the two groups (n = 7 healthy, 11 PASC). We also assessed knee extensor force and found that there was no significant difference in the muscle’s ability to produce force (**Fig 1C**, n = 7 healthy, 11 PASC). Subjects with PASC achieved on average 9.8% lower of their expected distance for 6-minute minute walk test based on age, sex and height (*p* = 0.0894) when compared to healthy control subjects (**Fig 1D**, n = 7 healthy, 11 PASC), which is a clinically relevant reduction in performance [27, 37, 38]. There was no difference in rectus femoris (**Fig 1E**) or vastus lateralis (VL) complex (**Fig 1F**) thickness when assessed via sonography (n = 7 healthy, 10 PASC). Immunohistochemical analysis of VL biopsies supported the ultrasound findings, with no significant differences in muscle fiber size (**Supplemental Fig 1A**). We also found no significant differences in fiber type composition (**Supplemental Fig 1B**). Although fiber type is coupled to oxidative capacity in healthy individuals, in some disease states these two variables are disconnected [39, 40]. We sought to gain insight into muscle mitochondrial activity using common histological methods: succinate dehydrogenase (SDH) histochemistry, which is a surrogate indicator of SDH enzyme activity. The intensity of SDH histochemistry was quantified as a percent of all fibers showing dark, intermediate, or light coloration. In the PASC group (n = 9), there was a significantly (*p* < 0.05) higher percentage of intermediate intensity fibers (**Supplemental Fig 1C**) than in the healthy group (n = 7), suggesting lower mitochondrial activity. Type I fibers trended (p < 0.1) towards lower levels of dark fibers with a trending (p < 0.1) higher level of intermediate fibers (**Supplemental Fig 1D**) in the PASC group. Type II fibers exhibited significantly higher levels of intermediate SDH stained fibers while having a significantly lower proportion of light SDH stained fibers **(Supplemental Fig 1E**).

**Figure 1.**
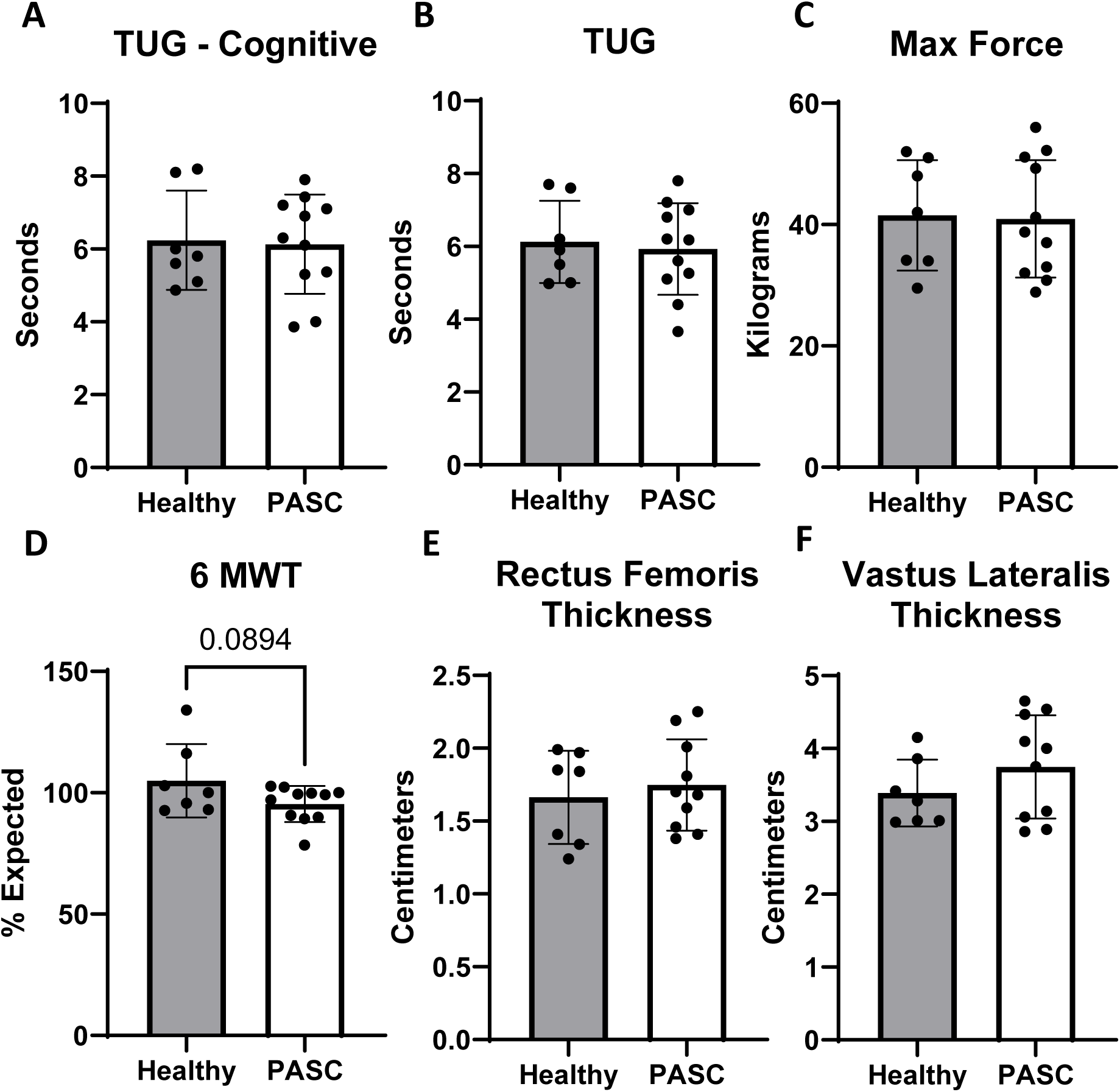
Muscle function testing and ultrasonography show no significant differences between PASC and healthy controls. **A and B.** Timed up and go (TUG) tests both with and without simultaneous cognitive testing in healthy (n=7) and PASC (n=11). **C**. Maximum force generated in knee extensors was not different between healthy (n=7) and PASC (n=11). **D.** Percent of expected 6-minute walk testing (6 MWT) was not statistically different but was trending (*p*=0.0894, n = 7 healthy, 11 PASC). **E and F**. Both rectus femoris and vastus lateralis thickness did not differ between healthy (n= 7) and PASC (n=10). Trending is defined as 0.05 < *p* ≤ 0.1.

### PASC participants have compromised skeletal muscle mitochondrial function

Several recent studies have linked PASC fatigue to reduced mitochondrial respiratory capacity in skeletal muscle [12, 41]. The observation that SDH activity differed in type II fibers prompted us to explore potential differences in mitochondrial respiration between healthy individuals and those with PASC. To investigate this, we used high resolution respirometry (HRR) to measure mitochondrial oxygen flux (*J*O2). State 3 respiration for complex I and combined complex I+II were significantly (*p* < 0.05) lower in PASC muscle compared to control (**Fig 2A-B**). Complex II respiration and maximal electron transfer (*ET*) capacity were not significantly different between groups (**Fig 2C-D**).

**Figure 2.**
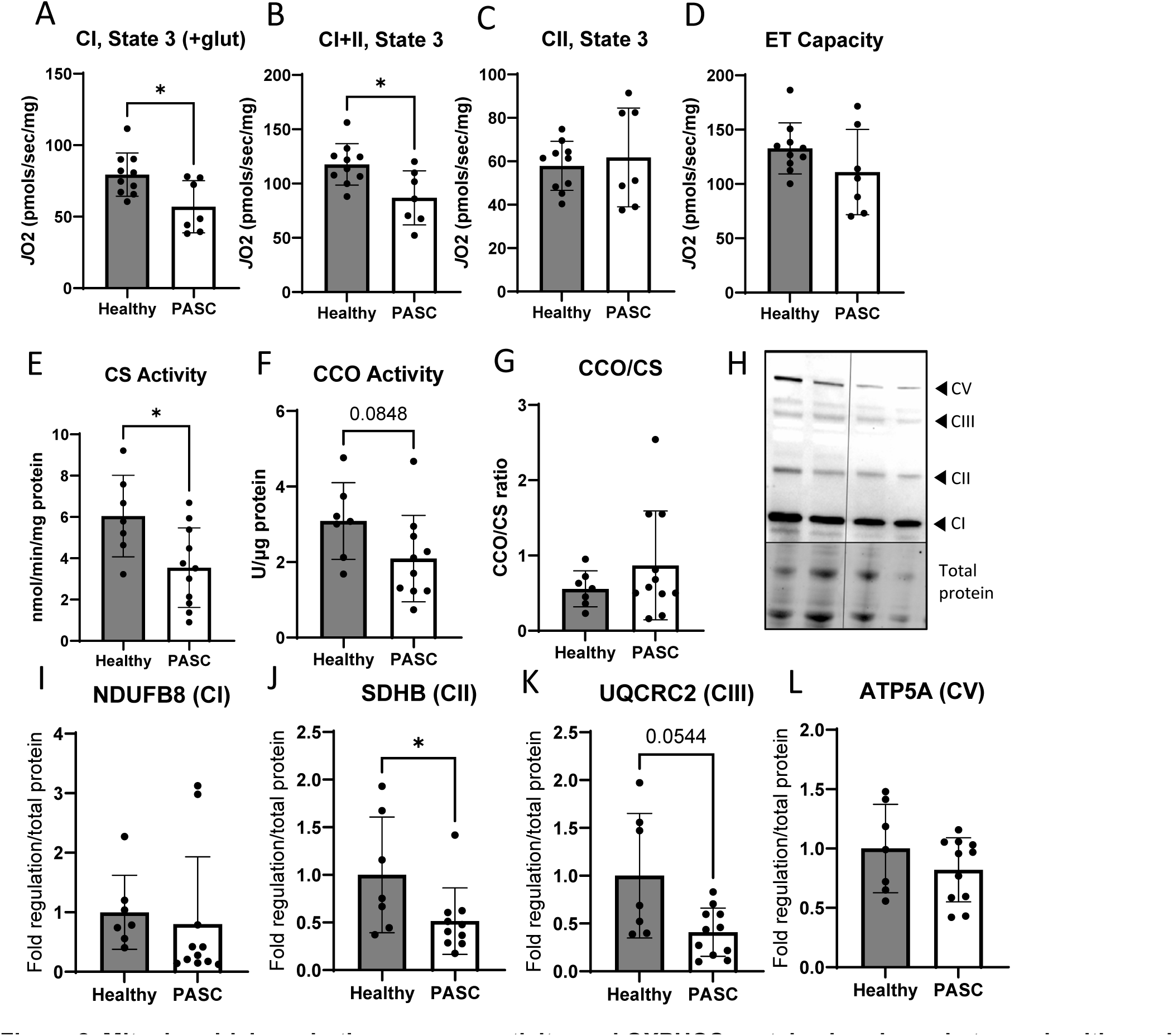
Mitochondrial respiration, enzyme activity, and OXPHOS protein abundance between healthy and PASC participants. **A -C.** Respiration is lower for complex I, state 3 + glutamate, and complex I+II, but unchanged in complex II, state 3 (n= 7 healthy, 11 PASC). **D.** Electron transport activity is not different between groups (n=10 healthy, 7 PASC). **E.** Citrate synthase (CS) activity is significantly different in PASC participants (n=7) when compared to healthy (n=10). **F.** Cytochrome C Oxidase (CCO) activity is trending but does not reach statistical significance. **G.** The ratio of CCO to CS is not significantly different. **H-L.** Western blot analysis and quantification of protein subunits in mitochondrial complex I (NDUFB8, **I**) complex II (SDHB, **J**), complex III (UQRCR2, **K**), and V (ATP5A, **L,** n= 7 healthy, 11 PASC). Data expressed as mean ± SD with symbols representing individual data points. * indicates *p* < 0.05 and trending is defined as 0.05 > *p* ≤ 0.1.

Citrate synthase activity and cytochrome C oxidase activity were further measured in tissue homogenates (**Fig 2E-F**). We found that citrate synthase (CS, a proxy for mitochondrial content) was significantly lower, but cytochrome C oxidase (CCO, a proxy for complex IV activity) did not reach statistical significance (*p* = 0.0848, n = 7 healthy, 11 PASC). The ratio of CCO to CS was not significantly different between groups (**Fig 2G**), suggesting that mainly mitochondrial content and not activity was lower in PASC compared to healthy muscle. Electron microscopy demonstrated several alterations in muscle mitochondrial morphology with occurrences of rupture as well as autophagosome localization near mitochondria (**Supplemental Fig 2**).

To determine if mitochondrial complex protein levels were stoichiometric, we performed western blot analysis (**Fig 2H**) of 4 key subunits of complex I (NDUFB8, **Fig 2I**), II (SDHB, **Fig 2J**), III (UQCRC2, **Fig 2K**), and V (ATP5A, **Fig 2L**). Quantification of those blots indicated complex II protein was significantly lower, and complex III approached significance (*p* = 0.0544, n = 7 healthy, 11 PASC). To determine if changes in mitochondrial content and function from SARs-CoV-2 infection were due to altered mitochondrial biogenesis, we measured synthesis rates of mitochondrial protein [42]. However, there were no statistical differences in the fractional synthesis rate (FSR) for muscle mitochondrial protein (**Supplemental Fig 3**), indicating that mitochondrial biogenesis was not impaired.

### Circulating sIL2R is significantly elevated in PASC participants and can directly impair mitochondrial function in muscle cells in vitro

Prolonged inflammatory response and T cell dysfunction have been linked to PASC fatigue, and many inflammatory markers are reported to be elevated systemically with PASC [43–45]. To assess circulating markers in our participants, we quantified 65 circulating factors (Human Immune Monitoring Panel) in the serum of PASC and healthy participants (**Fig 3**, n = 8 healthy, 6 PASC). We found sIL2R was the most highly abundant circulating inflammatory marker, with the largest fold difference between healthy and PASC (*p* <0.05), followed by IL-3, IL-22, IL-21, and IL-31. Considering the most recent study linking T cell dysfunction to muscle-associated PASC fatigue [41], we hypothesized that sIL2R directly affects mitochondrial respiration.

**Figure 3.**
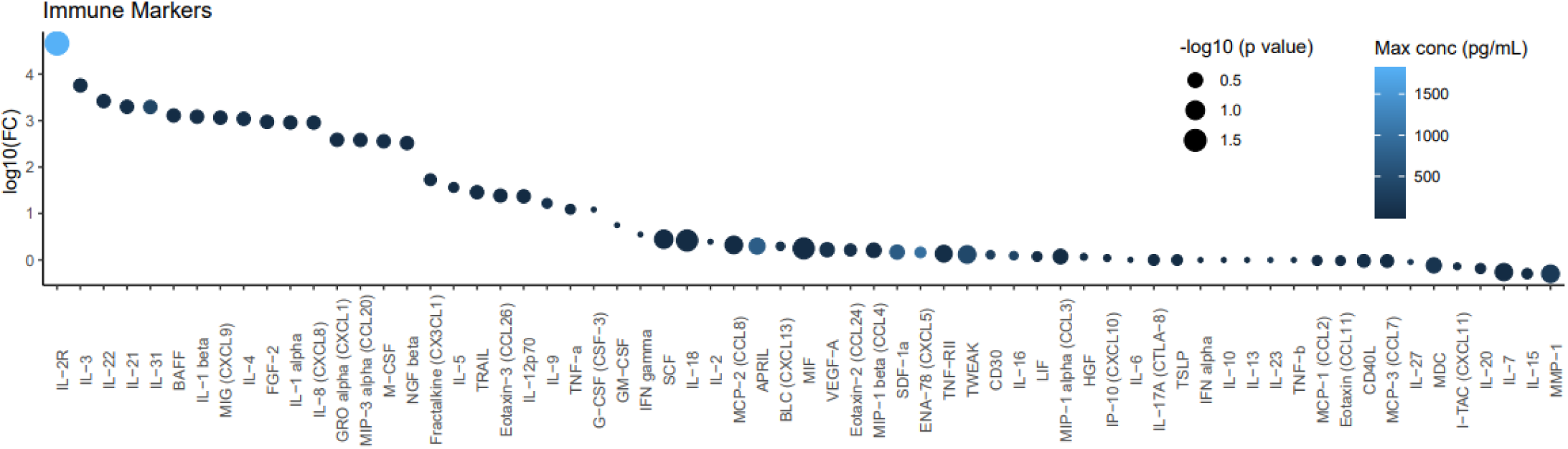
Inflammatory markers in PASC. Dot plot with the Y axis indicating log10 fold change of inflammatory markers in PASC patients when compared to healthy. Dot size negatively scales with *p* value, and color indicates the markers’s highest (Healthy or PASC) concentration in serum (pg/ml). N = 8 healthy, 6 PASC.

To assess if sIL2R has an inhibitory effect on mitochondrial respiration, we treated C_2_C_12_ myotubes with human recombinant sIL2R or IL-2 for 24 hours and used a Seahorse Extracellular Flux Analyzer to assess oxygen consumption rate (OCR) (**Fig 4A**, n = 4). We found that basal respiration (**Fig 4B**) was unchanged, but maximal respiration (**Fig 4C**) was significantly reduced after treatment with sIL2R when compared to vehicle (PBS). Administration of IL-2 had no significant effect on OCR when compared to vehicle (**Supplemental Fig 4**), suggesting that the mechanism of action for sIL2R is distinct from known IL-2 pathways described for T cells. We assessed CCO activity (**Fig 4D**, n = 3-4) and found that myotubes treated with sIL2R have significantly lower complex-IV activity. We performed a western blot analysis (**Fig 4E**, n = 3) and found that complex III subunit protein (UQCRC2) was significantly lower with sIL2R administration (**Fig 4H**). Mitochondrial proteins ATP5A (CV), SDHB (CII), and NDUFB8 (CI) were trending to be lower but did not reach statistical significance (**Fig 4F, G, and I**).

**Figure 4.**
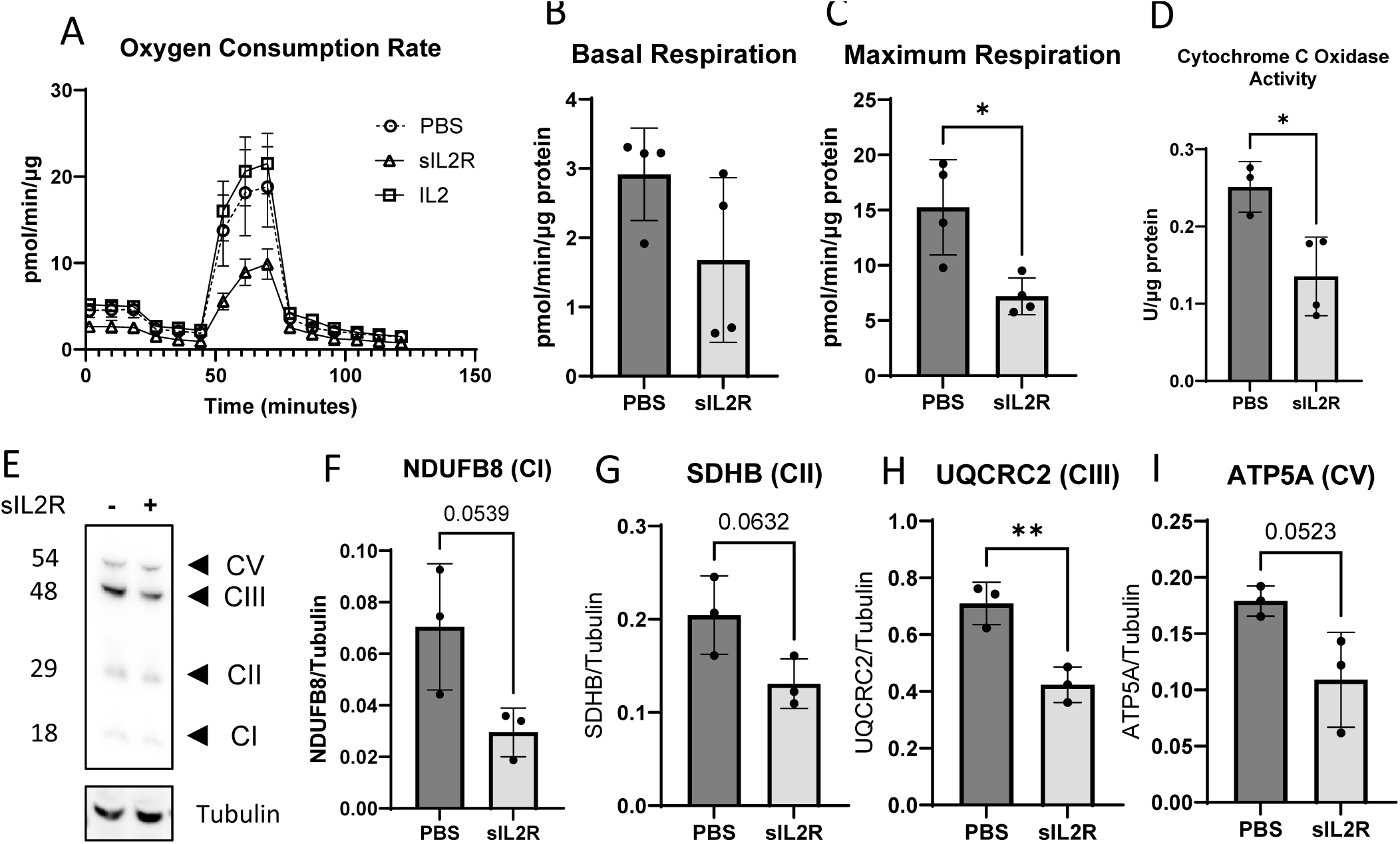
IL-2R application to myotubes. **A.** Oxygen consumption is reduced in myotubes after sIL2R administration but is not affected by IL-2 administration (n=4). **B**. Basal respiration was not different between PBS and sIL2R treatment. **C**. Maximal respiration was lower in sIL2R cells. **D.** Cytochrome C oxidase activity was significantly different between PBS (n=3) and sIL2R (n=4). **E**. Western blot analysis of C_2_C_12_ myotube lysates showing complex V (CV) protein ATP5A, complex III (CIII) protein UQCRC2, complex II protein SDHB, and complex I protein NDUFB8 with and without sIL2R (n=3). **F – I .** Quantification of protein subunits in mitochondrial complex I (NDUFB8, **F**) complex II (SDHB, **G**), complex III (UQRCR2, **H**), and V (ATP5A, **I,** n=3). Data expressed as mean ± SD. ** indicates *p*< 0.01, * indicates *p* < 0.05, and trending is defined as 0.05 < *p* ≤ 0.1.

## DISCUSSION

In this study we investigated skeletal muscle and systemic factors potentially responsible for persistent fatigue in PASC participants. Despite reporting chronic fatigue and achieving 10% less expected 6-minute walking distance compared to controls, PASC participants showed no apparent differences in muscle size or strength. However, they exhibited significant impairment of mitochondrial respiration across multiple components of the respiratory chain. To elucidate the mechanism behind this skeletal muscle mitochondrial respiratory deficiency, we assessed circulating factors -soluble interleukin-2 receptor (sIL2R) emerged as the most significantly elevated factor in PASC patients. Notably, *in vitro* treatment of myotubes with sIL2R impaired respiration, suggesting a direct link between this circulating factor and respiratory capacity.

Among the elevated immune markers in our PASC cohort, sIL2R (CD25) showed the highest expression and greatest difference from healthy controls. Elevated sIL2R levels have been correlated to muscle wasting like sarcopenia [46] and cachexia [45] and, in COVID, sIL2R was predictive of respiratory failure [44] and ICU outcomes [47]. Our findings extend these observations, proposing a mechanism by which sIL2R might contribute to worsened outcomes and persistent fatigue after recovery from acute COVID. Our *in vitro* data suggest that IL-2 alone does not significantly affect muscle cell mitochondrial respiration, whereas sIL2R can inhibit mitochondrial OXPHOS even at relatively low levels. Therefore, we posit that PASC fatigue is partially attributable to elevated systemic sIL2R inhibiting mitochondrial oxygen consumption in muscle cells.

IL-2R is a trimeric protein complex that is located on the plasma membrane and is primarily found on T cells; thus, most of current understanding of IL2R function is based on T cell biology and IL-2 signaling [48, 49]. IL-2 binds weakly to monomers of IL-2R (α chain), and heterodimers of the β and ɣ chain[49, 50]. However, as a trimer, IL-2 first binds the α chain causing a conformational change to IL-2 which increases its affinity for IL-2Rβ and IL-2Rɣ by one thousand-fold [50, 51]. The β chain is complexed with JAK1, and the ɣ chain is complexed with JAK3, tyrosine kinases. When IL-2R is activated by binding IL-2, three signaling pathways are initiated: MAP kinase, PI3K, and JAK/STAT. Although we have not found evidence to suggest that IL-2R is expressed in skeletal muscle, its expression in vascular smooth muscle cells (VSMC) has been shown to aid in VSMC maintenance during aging [52]. Interestingly, activated and proliferating VSMCs express lower levels of CD25 than quiescent VSMCs [52]. Arumugam et al suggest a delicate balance between low IL-2/IL2R signaling increasing proliferation via AKT, but higher levels activating FoxO3a and signaling survival/maintenance [52]. It is possible that such a mechanism is in place in skeletal muscle and that sIL2R is correlated with muscle impairment due high IL-2/IL2R signaling activating pro-maintenance systems and dampening proliferative signaling. However, this pathway does not explain our findings that direct administration of sIL2R, is capable of eliciting mitochondrial alterations *in vitro* and further experimentation is needed to elucidate the mechanisms involved in the action of IL2R.

Our experiments revealed that sIL2R administration in myotubes did not affect basal respiration, but had decreased maximal respiration. In addition, mitochondrial content was decreased in muscles from patients with PACS compared to controls. There findings could explain post-exertional malaise in PASC patients despite seemingly normal baseline muscle function. It suggests that mitochondria may function adequately under resting conditions but fail to meet increased energy demands during exertion [7, 11, 41, 53]. The link between post-viral fatigue and mitochondrial abnormalities in muscle has been recognized since the 1990’s [54]. While our findings stem from COVID participants, they may have broader implications for other viral infections and post-viral care. Furthermore, although we focused on skeletal muscle, sIL2R might potentially impair mitochondrial respiration in other tissues, such as the central nervous system. Future research could explore whether sIL2R contributes to cognitive symptoms like “brain fog” in PASC patients.

It would be valuable to address the mechanistic aspects of soluble interleukin-2 receptor (sIL2R) action on mitochondria in muscle tissue. While the exact pathway remains speculative, exploring potential mechanisms could provide a framework for future investigations. One possibility is that sIL2R interacts with membrane-bound receptors on muscle cells, initiating a signaling cascade that ultimately affects mitochondrial function. One such possibility is the β1-integrin, which may interact with extracellular IL-2R to mediate intracellular signaling that regulates myoblast proliferation and differentiation [55]. Another possibility is a mechanism of *trans*-signaling reminiscent of soluble IL-6R (sIL-6R), where the sIL-6R-IL-6 complex binds to gp130 to mediate intracellular potentiation of proinflammatory pathways [56]. Alternatively, sIL2R might cross the muscle cell membrane through yet-unidentified transport mechanisms, directly influencing intracellular processes. The downstream effects on mitochondria could involve alterations in mitochondrial turnover, oxidative phosphorylation, or mitochondrial dynamics. Elucidating these potential mechanisms would not only enhance our understanding of sIL2R’s role in muscle physiology but also provide insights into novel therapeutic targets for other tissues. Further research is needed to delineate the precise molecular pathways involved in this intriguing interaction between sIL2R and muscle mitochondria.

Some limitations exist for this study. The availability of samples with known negative COVID status is limited, but we made every effort to optimize our research within these constraints. We included both pre-and post-pandemic controls samples, which resulted in higher than optimal levels of variability in age and BMI. Variability can act as a potential confounding factor, and therefore the significance of our results, despite high variability, suggests a strong biological effect. It is currently unclear if the alterations in OXPHOS are due to total number of mitochondria or due to decreased capacity per mitochondria. Our data shows that CS activity is less in both *in vivo* and *in vitro* indicating an overall reduction in mitochondrial content and this data is in agreement with the anecdotal TEM findings (n=1), but a more in-depth analysis is required to confirm lower mitochondrial content. In conclusion, our study provides new insights into the physiological basis of PASC fatigue, highlighting the role of sIL2R in mitochondrial dysfunction. These findings open avenues for potential therapeutic interventions and underscore the need for comprehensive post-viral care strategies.

## Contributions

LPB analyzed data, performed experiments, and took the lead in preparing the manuscript and figures. DEL collected blood and performed functional experiments. PAK and AGK collected muscle biopsies. JJ, AAM, KK, HV, DW, and CL performed experiments and analyzed results. JLS, PS PAK, CF, EED provided mentorship, and provided guidance throughout. AI performed experiments, analyzed results and helped prepare the manuscript. KPM conceived the project, supervised, helped write and oversaw the overall direction and planning. YW designed experiments, helped write the manuscript, and guided the direction of the research. All authors provided critical feedback and contributed to the final version of the manuscript.

## Data Availability

All data produced in the present study are available upon reasonable request to the authors.

## Acknowledgements

The authors would like to thank Sandra Rigsby for her help with the fractional protein synthesis experiment.

