## Supplemental figures for "Soluble IL-2R contributes to impaired muscle cell mitochondrial respiration in fatigued individuals with post-acute sequelae of COVID" (from another version of this preprint: version 1, 2024-08-15)

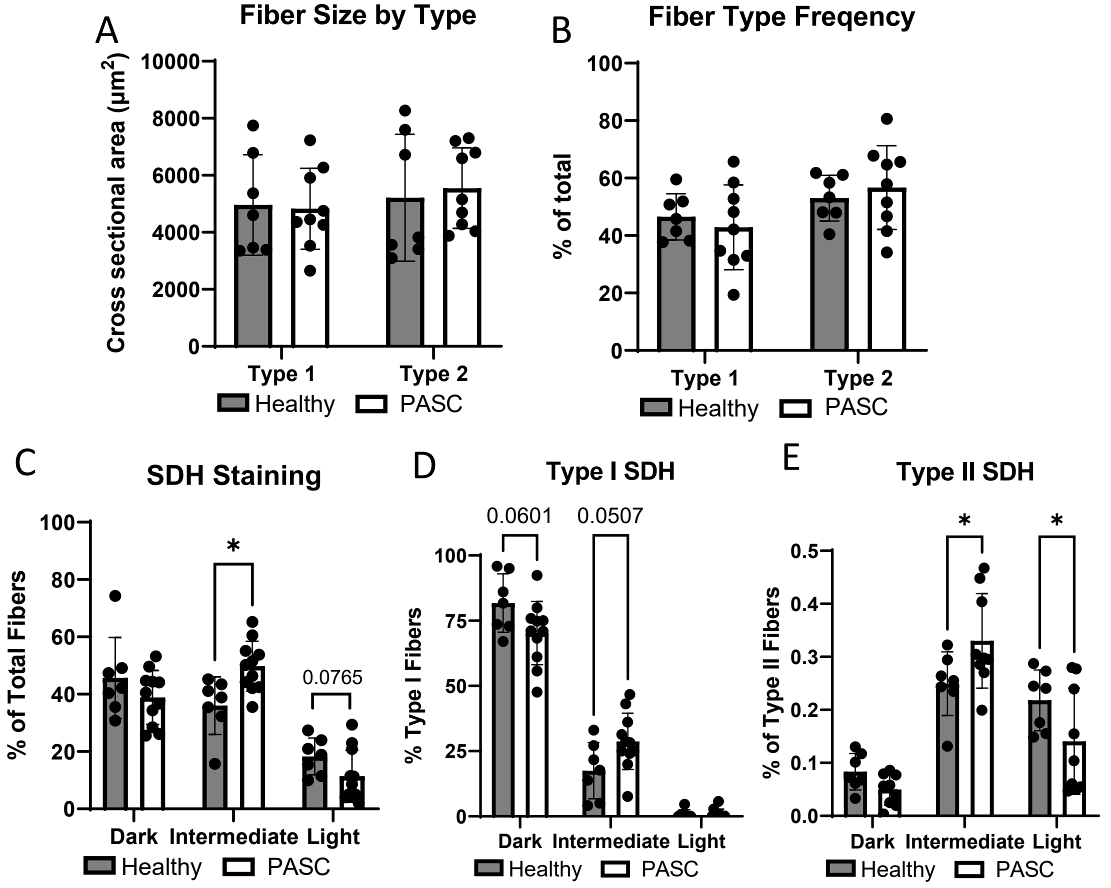


**Supplemental Figure 1. Histological indicators of altered muscle fiber energetics within vastus lateralis biopsies from participants with PASC compared to healthy individuals.** Quantification of muscle fiber size, fiber type, and mitochondrial activity/SDH within biopsies from PASC (n=9) participants compared to healthy adults (n=7). Data expressed as mean ± SD. **A.** Fiber type specific CSA from PASC participants, compared to healthy. **B.** Frequency of fibers expressing Type I, or Type II MyHC within muscle from PASC versus healthy adults. **D.** Quantification of muscle fiber SDH histochemistry in muscle showing a higher number of intermediate fibers in PASC. **D and E.** Quantification of SDH histochemistry within individual fiber types. **D.** Type 1, **E.** Type 2. N = 7 healthy, 9 PASC. * indicates $p < 0.05$, and trending is defined as $0.05 < p \leq 0.1$.

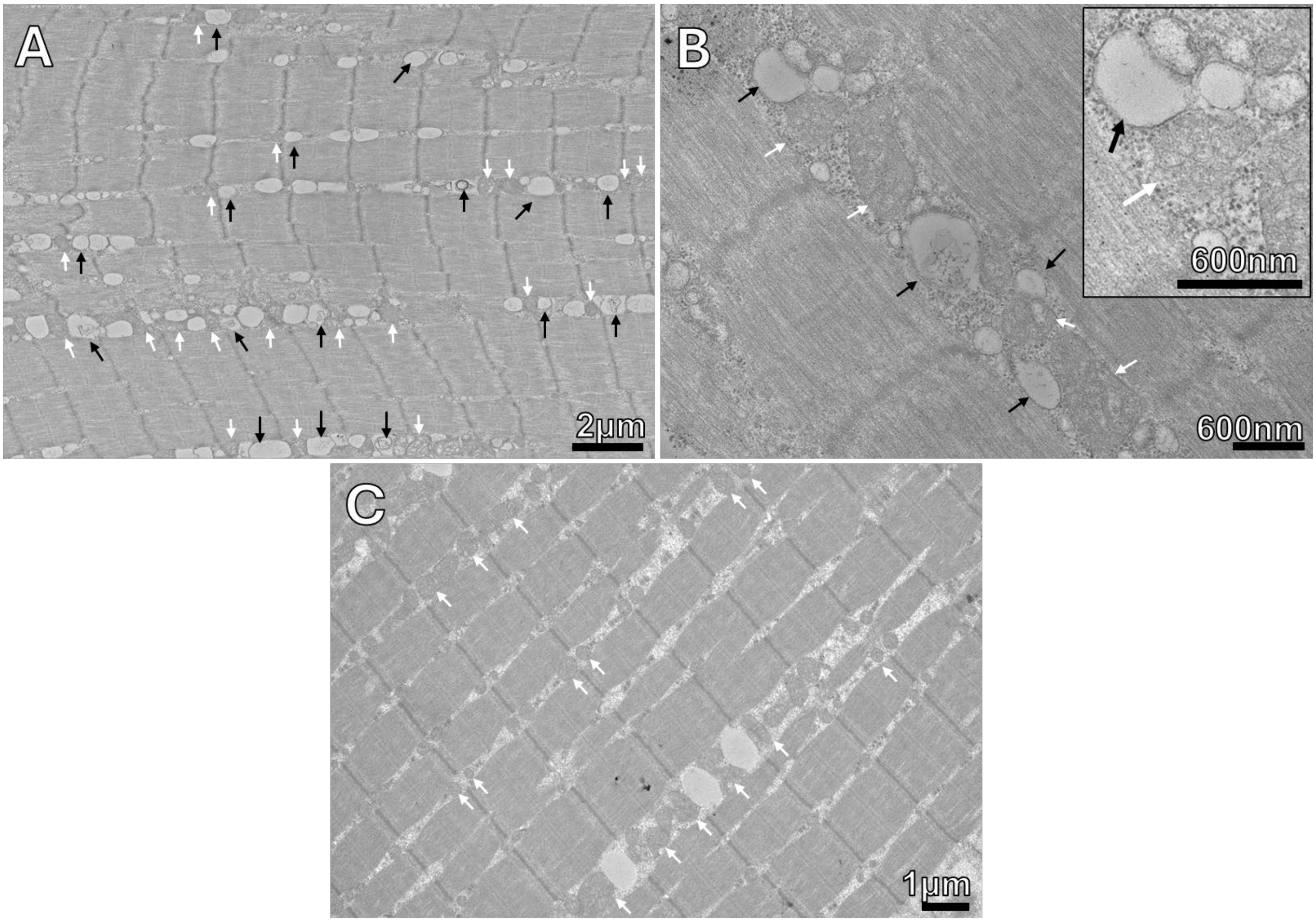


**Supplemental Figure 2. Increased autophagosomes localized near mitochondria.** Pannels **A** and **B** show representative micrographs of a PASC participant's skeletal muscle sarcomere and mitochondrial features at 2µm and 600nm. Compared to control muscle (**C**) PASC displayed fewer mitochondrial pairs at Z-discs and increased autophagosome (black arrows) localization near mitochondria (white arrows).

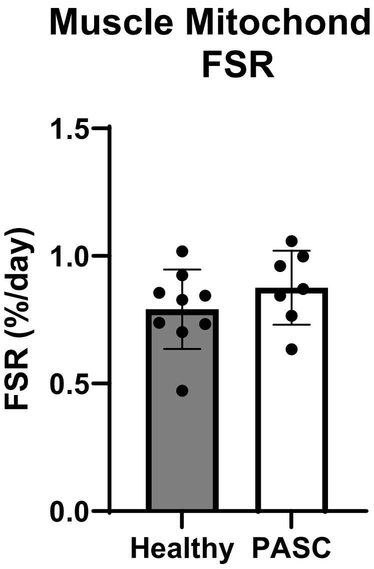


**Supplemental Figure 3. Muscle mitochondria fractional protein synthesis (FSR).** No differences in FSR (%/day) n = 9 healthy, 7 PASC. p = 0.2523 (Mann-Whitney U test)

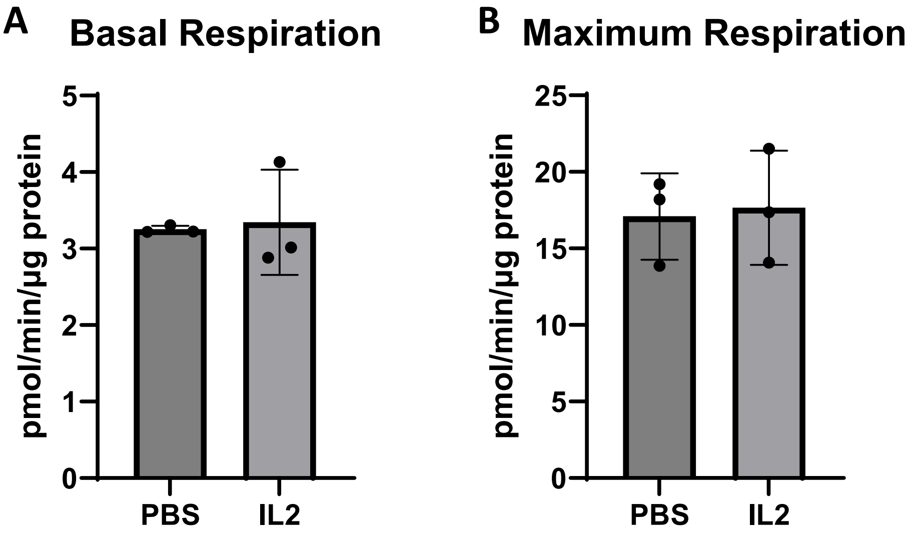


**Supplemental Figure 4. Cellular respiration after administration of IL2 on C2C12 myotubes.** Basal respiration (**A**) and maximal respiration (**B**) is unchanged after IL2 is administered on myotubes. N=3.
